# Variability in Radiotherapy Outcomes Across Cancer Types: A Comparative Study of Glioblastoma Multiforme and Low-Grade Gliomas

**DOI:** 10.1101/2024.07.04.24309952

**Authors:** Alexander Veviorskiy, Garik V Mkrtchyan, Andreyan N Osipov, Ivan V Ozerov, Alex Aliper, Alex Zhavoronkov, Morten Scheibye-Knudsen

## Abstract

Radiotherapy is a crucial treatment option for various cancers. However, the results of radiotherapy can vary widely across different cancer types and even among patients with the same type of cancer. This variability presents a major challenge in optimizing treatment strategies and improving patient survival. Here, we collected radiotherapy phenotype and expression data from 32 TCGA cancer datasets and performed overall survival analysis for 32 cancer types. Additionally, we conducted a signalling pathway enrichment analysis to identify key pathways involved in radiotherapy resistance and sensitivity. Our findings show that radiotherapy improves survival outcomes in certain cancer types, such as GBM, while worsening outcomes in others, such as LGG. Next, we focused on exploring the differences in radiotherapy outcomes between GBM and LGG, focusing on the molecular mechanisms contributing to these variations. The differential regulation of pathways related to programmed cell death, DNA repair, telomere maintenance, chromosome condensation, antiviral responses, and interferon signaling between GBM and LGG patients may elucidate the underlying reasons for these observed differences. These insights underscore the importance of personalized treatment approaches and the need for further research to improve radiotherapy outcomes in cancer patients.

## Introduction

Radiotherapy plays a fundamental role in the treatment of cancer. Even though radiotherapy is widely used, its outcomes can vary significantly depending on the cancer type. It was observed that GBM patients who received radiotherapy are better overall survivors than those who didn’t receive radiotherapy [1]. Conversely, the opposite effect was observed with LGG patients [2]. Tumor heterogeneity is a major factor that affects radiotherapy response rates, even among patients diagnosed with the same tumor type [3]. The importance of studying variability in radiotherapy outcomes across cancer types lies in the complex interactions between treatment, anatomical, and patient-related variables. These interactions can significantly influence treatment efficacy and patient prognosis [4]. Additionally, the use of radiotherapy varies significantly across different cancer diagnoses, and understanding these variations can help improve treatment strategies [5]. GBM and LGG are particularly interesting to study together because GBM often originates from a preexisting LGG, representing a progression from a lower-grade to a higher-grade malignancy [6]. This progression is associated with significant changes in gene expression profiles [7], which may underlie the differences in radiotherapy outcomes observed between these two cancer types. Understanding these differences is crucial, as it can inform personalized treatment strategies and improve survival outcomes.

Recent advancements in radiotherapy for GBM and LGG include the exploration of various targeted therapies, novel radiotherapy approaches, and immunotherapies. For instance, vaccine strategies have shown promising results in early-phase clinical studies [8]. Particle irradiation and dose escalation strategies, including modern molecular imaging, are being evaluated for their long-term outcomes [9]. Intraoperative radiotherapy (IORT) is being explored to sterilize the margins from persistent tumor cells and bridge the therapeutic gap between surgery and radiochemotherapy [10]. Immunotherapy is another promising modality, with radiotherapy potentially enhancing the effect of immunotherapy through various mechanisms [11].

Despite these advancements, the survival rate for GBM patients has not significantly improved, indicating a need for novel anti-target therapies that could be used in conjunction with standard radiochemotherapy approaches [12]. Radiotherapy resistance is frequently observed in GBM patients and is a major cause of the high mortality rate [13]. This resistance is often multifactorial and heterogeneous, associated with the recurrence of GBM after surgery [14]. For LGG, while radiotherapy is effective, it is associated with neurological, cognitive, and endocrinological morbidity, prompting the use of chemotherapy regimens aimed at delaying radiotherapy, especially in younger children [15].

In response to these challenges, our study aims to investigate the variability in radiotherapy outcomes between GBM and LGG patients. We collected radiotherapy phenotype data for 32 TCGA cancer datasets and performed an overall survival analysis for 32 cancer types. We also conducted a signaling pathway enrichment analysis to uncover the underlying biological processes contributing to the observed differences in radiotherapy outcomes for GBM and LGG cancer patients. The differential regulation of pathways related to programmed cell death, DNA repair, telomere maintenance, chromosome condensation, antiviral responses, and interferon signaling was observed. The findings could have a significant impact on personalized treatment approaches and novel co-treatment approaches with RT.

## Materials and Methods

### Data Collection and Differential Expression Analysis

Gene expression and patient phenotype data for 32 TCGA cancers were collected from the UCSC Xena database [16]. FPKM-UQ expression data obtained from tumor samples were uploaded into PandOmics [17] and pre-processed according to the PandaOmics pipeline. Differential expression analysis was performed using the limma package for TCGA-GBM and TCGA-LGG datasets, comparing patients who received radiotherapy to those who did not receive radiotherapy. Each dataset was processed according to standard protocols. The obtained gene-wise p-values were corrected using the Benjamini–Hochberg procedure. Differential expression results were later used for gene set enrichment analysis.

### Overall survival analysis of IR-treated and IR-untreated cancer patients

Survival analysis was conducted in PandaOmics using the KaplanMeierFitter function from the lifelines Python package. Patients were divided into two groups: those who received radiotherapy and those who did not receive radiotherapy. Only patients with available expression data were included in the analysis. Briefly, 32 TCGA cancers were analyzed, and survival analysis was performed between the two described groups of patients. The log-rank test was used to calculate statistical significance. The significance of survival outcomes was plotted on a heatmap and colored red if radiotherapy increased survival outcomes and blue if the application of radiotherapy decreased survival outcomes. Non-significant results were colored white. Combined survival plots for patients who received and did not receive radiotherapy across all TCGA cancers were plotted using the matplotlib package.

### Signaling pathway enrichment analysis

Pathway enrichment analysis was performed using the gseapy package with the enrichr() function, following standard protocols. The Reactome database was selected as the source of gene sets from the gseapy internal library for signaling pathway enrichment analysis. Genes that were significantly up-regulated in TCGA-GBM and simultaneously down-regulated in TCGA-LGG, and vice versa, were used as input for the pathway enrichment analysis. The top 20 significantly perturbed signaling pathways were visualized on a dot plot using the gseapy.plot function.

### Paper draft preparation

The draft of this paper was generated using DORA, Insilico Medicine's LLM-based paper drafting assistant. Draft Outline Research Assistant (DORA) is designed to streamline the process of publication creation, making it faster and simpler. The process of paper generation is curated by over 30 AI agents, powered by Large Language Models (LLMs), and integrated with internal and other curated databases, to assist in generating high-quality scientific papers. Each agent employs Retrieval-Augmented Generation (RAG) to perform comprehensive data collection and analysis, reduce the probability of hallucinations, and provide relevant PubMed links to make the generation of the paper more transparent. Followed by generation, the draft was manually curated and extended by the authors.

## Results

### Patient and Tumor Characteristics in TCGA Cancers

Radiotherapy phenotype data were collected for 32 TCGA cancer datasets from the UCSC Xena database. TCGA-LAML was excluded from the analysis since there were no patients who received radiotherapy. The total number of samples varies between 1,194 and 45 for TCGA-BRCA and TCGA-CHOL, respectively. Similarly, the percentage of patients who received radiotherapy varies between 82% and 0% for TCGA-GBM and TCGA-KIRC/TCGA-CHOL/TCGA-KICH/TCGA-KIRP, respectively. It was noted that TCGA-GBM and TCGA-LGG are the cancers with the highest percentage of patients who received radiotherapy, at 82% and 54%, respectively (Figure 1).

**Figure 1.**
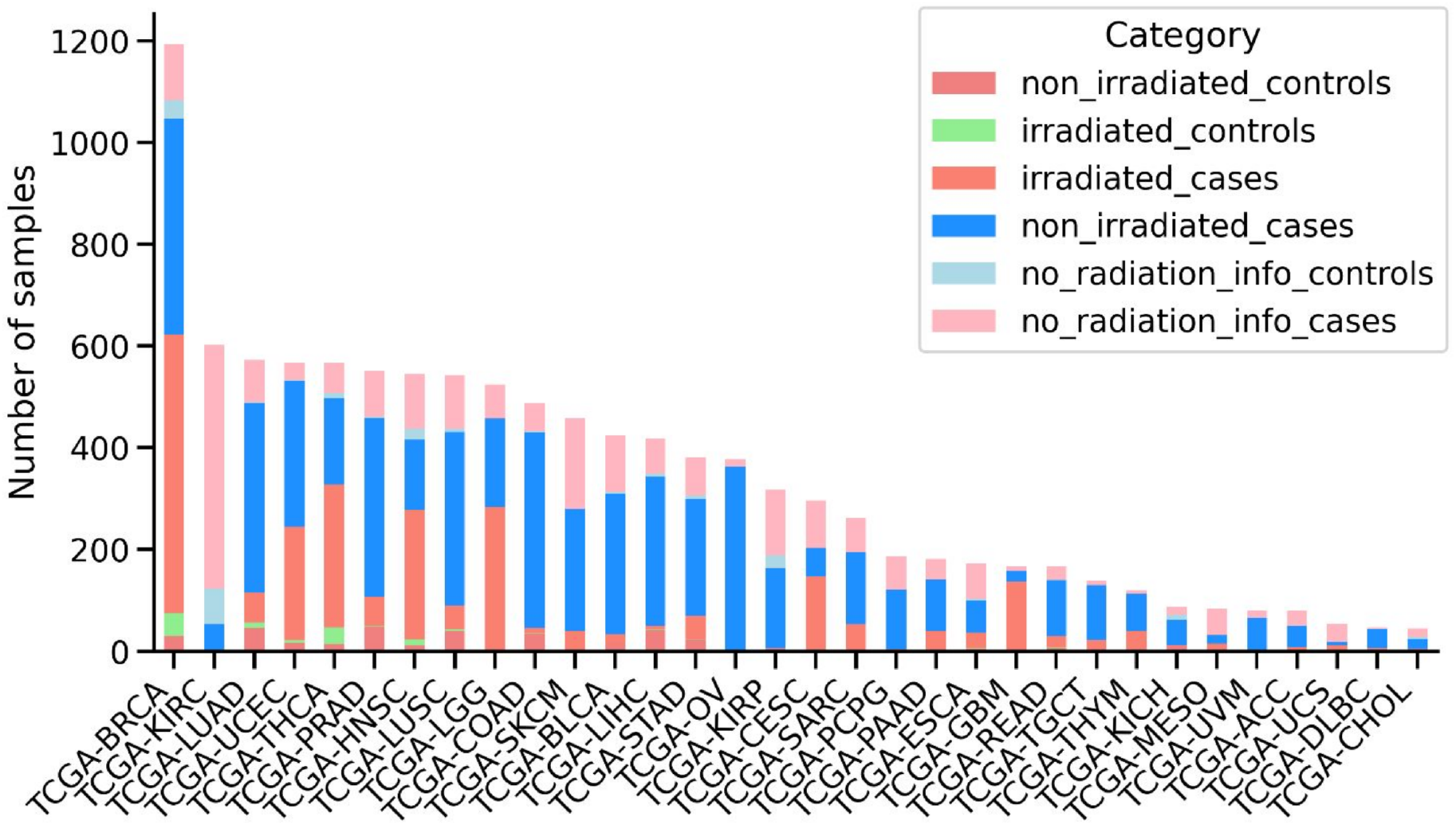
Overview of the patient’s samples downloaded from TCGA. Number of samples for each cancer type is shown, sorted by the total number of samples. Stacked bars are colored according to the sample category, including non-irradiated control samples, irradiated control samples, irradiated case samples, non-irradiated case samples, control samples without information about radiotherapy and case samples without information about radiotherapy.

### Overall survival analysis of patients who received and did not receive radiotherapy

To study whether radiotherapy treatment could be used as a trait capable of stratifying cancer patients with different outcomes, we performed an overall survival analysis for 32 cancer types from the TCGA dataset. The overall survival analysis was conducted for patients who received radiotherapy and for those who did not receive radiotherapy (Figure 2, A). In some cases, radiotherapy can improve survival outcomes, while in others, it worsens them. For example, it was found that patients with GBM, BRCA, READ, UCEC, STAD, and HNSC who received radiotherapy lived longer compared to patients who did not receive radiotherapy. Conversely, it was noted that patients with UVM, LUAD, and LGG who received radiotherapy had shorter survival compared to those who did not receive radiotherapy (Figure 2, B). This observation led us to focus on the differences between GBM and LGG patients who received and did not receive radiotherapy, since GBM can develop from LGG.

**Figure 2.**
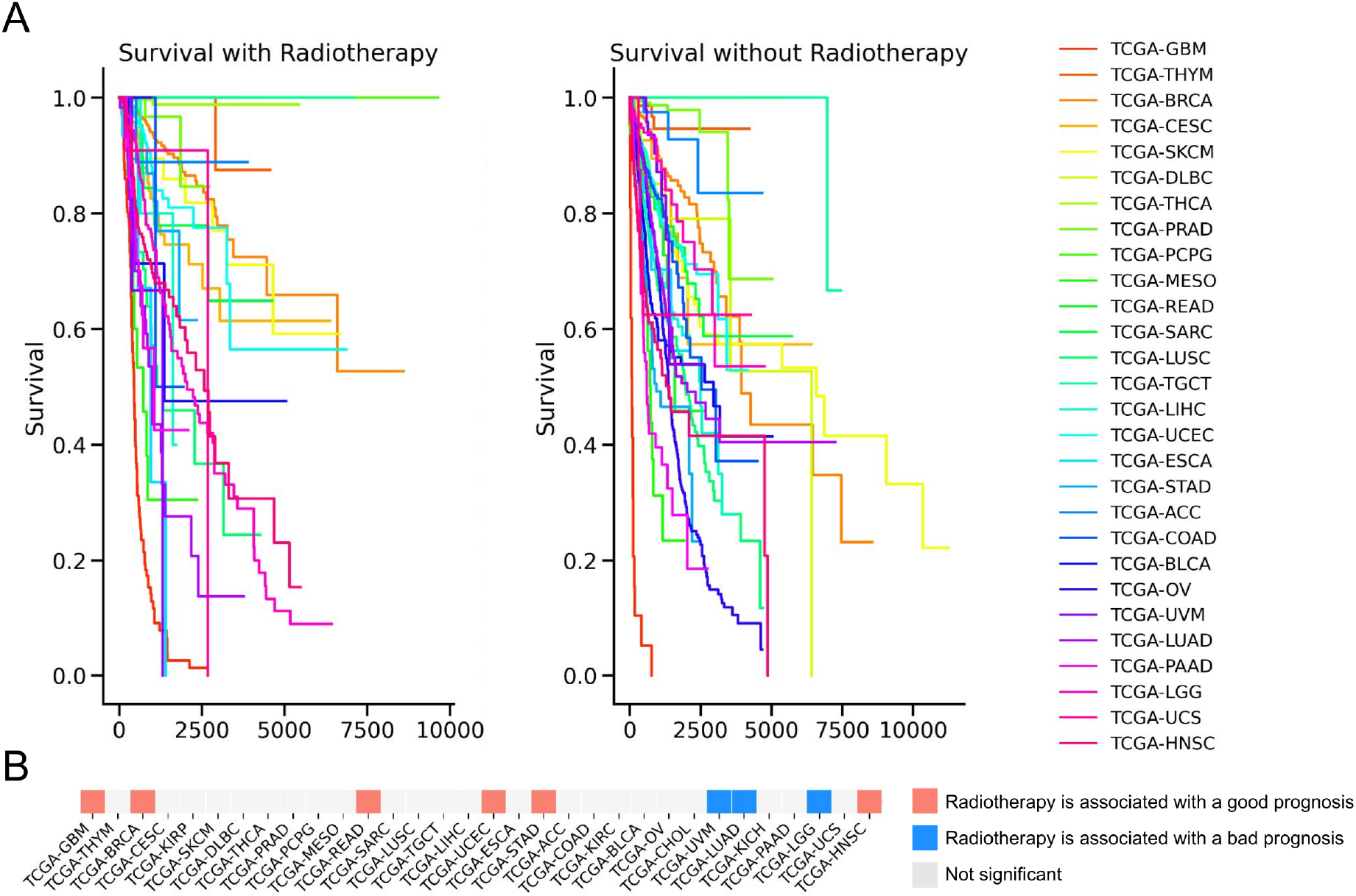
Survival analysis across 22 TCGA cancers. A, Survival curves for patients who received radiotherapy and those who didn’t presented on a Kaplan-Meier plot. B, The significance of survival results is plotted on a heatmap and colored red if radiotherapy increased survival outcomes and blue if the application of radiotherapy decreased survival outcomes. Non-significant results were colored white (p-value > 0.05). The log-rank test was used to calculate statistical significance.

### Signaling pathway enrichment analysis of GBM and LGG patients

In order to find differences between GBM and LGG patients who were exposed to IR and who weren’t on the transcriptomic level, we collected gene expression data for those patients. Then we created comparisons between patients who received radiotherapy and those who did not, and calculated differentially expressed genes. After that, we obtained a list of genes that were significantly down-regulated in GBM patients and at the same time significantly up-regulated in LGG patients, and vice versa. Those gene lists were used for pathway enrichment analysis to uncover the underlying biological processes contributing to the observed differences in radiotherapy outcomes (Figure 3). It was noted that pathways related to programmed cell death and DNA repair, such as "Diseases Of Programmed Cell Death" and "Base-Excision Repair," were down-regulated in GBM but up-regulated in LGG. This suggests a potential mechanism by which LGG cells might be more susceptible to radiotherapy-induced damage, whereas GBM cells might evade such damage. It was also observed that pathways involved in telomere maintenance and chromosome condensation, such as "Packaging Of Telomere Ends" and "Condensation Of Prophase Chromosomes," were differentially regulated, indicating a possible role in the differential radiotherapy outcomes between GBM and LGG (Figure 3, A). Moreover, pathways associated with antiviral responses and interferon signaling, including "ISG15 Antiviral Mechanism" and "Interferon Signaling," were up-regulated in GBM and down-regulated in LGG. This could imply an enhanced immune response in GBM, potentially contributing to its resistance to radiotherapy. Finally, pathways related to gene expression and protein metabolism, such as "Gene Expression (Transcription)" and "Metabolism Of Proteins," were up-regulated in GBM and down-regulated in LGG, suggesting a higher metabolic and transcriptional activity in GBM that might support its aggressive nature and resistance to treatment (Figure 3, B).

**Figure 3.**
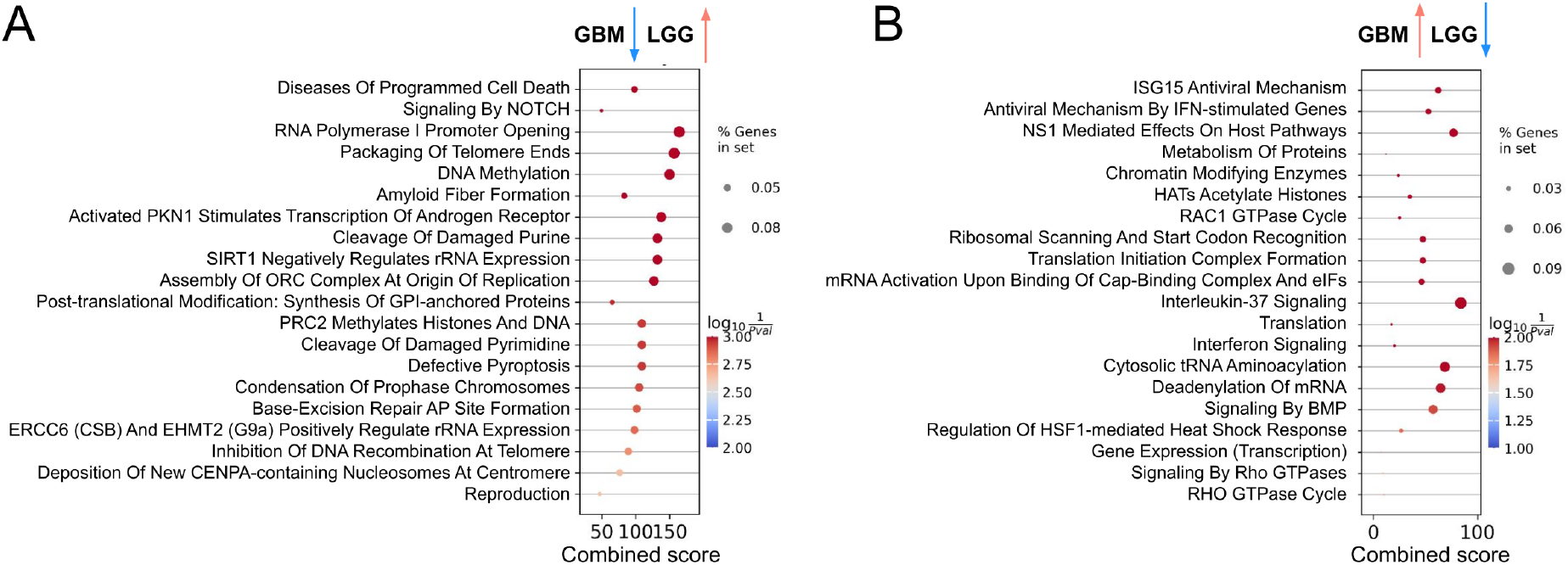
Signaling pathway enrichment analysis results. A, Signaling pathways enriched with genes significantly down-regulated in GBM patients and at the same time significantly up-regulated in LGG patients. B, Signaling pathways enriched with genes significantly up-regulated in GBM patients and at the same time significantly down-regulated in LGG patients.

## Discussion

The study presented here shows the variability in radiotherapy outcomes across different cancer types, with a specific focus on glioblastoma multiforme (GBM) and low-grade gliomas (LGG). One of the unique aspects of this study is its focus on the differential radiotherapy outcomes between GBM and LGG. While radiotherapy remains a fundamental treatment for GBM, the high resistance to treatment and subsequent poor prognosis emphasize the need for novel therapeutic strategies [18]. Conversely, LGG patients often show a more favorable response to radiotherapy, which can be predicted by MRI evaluations post-treatment [19]. The findings from this study have the potential to impact clinical practices and treatment protocols by providing a more detailed understanding of how radiotherapy should be tailored to individual patients based on their specific cancer type.

Radiotherapy was found to improve survival outcomes in GBM while worsening outcomes in LGG. GBM and LGG, despite their differences, share a common lineage, with GBM often developing from LGG [6]. This study addresses a critical gap in the existing literature by providing a detailed comparison of radiotherapy outcomes in these two cancer types. The differential regulation of pathways related to programmed cell death, DNA repair, telomere maintenance, chromosome condensation, antiviral responses, and interferon signaling between IR-exposed and not non-IR-exposed GBM and LGG patients may explain the underlying reasons for these observed differences. For instance, the down-regulation of DNA repair pathways in GBM suggests a mechanism for radiotherapy resistance, while their up-regulation in LGG indicates a higher susceptibility to radiotherapy-induced damage. The use of radiotherapy in combination with temozolomide has been shown to improve survival rates in GBM patients [20]. However, radiotherapy resistance remains a significant challenge, often leading to poor outcomes [13]. In LGG, the timing and dosage of radiotherapy are crucial factors that can influence survival outcomes [21]. Given the up-regulation of DNA repair pathways in LGG, combining radiotherapy with DNA repair inhibitors could make LGG cells more vulnerable to treatment. For example, using PARP inhibitors could enhance the effectiveness of radiotherapy [22]. On the other hand, the down-regulation of antiviral and interferon signaling pathways in LGG indicates a less active immune environment. Combining radiotherapy with immune modulators, such as interferon therapy or immune checkpoint inhibitors, could boost the immune response against LGG cells and improve treatment outcomes [23]. The results of this study align with these findings, further emphasizing the importance of personalized treatment approaches.

In conclusion, this study provides valuable insights into the variability in radiotherapy outcomes across different cancer types, with a specific focus on GBM and LGG. The identification of key pathways involved in radiotherapy resistance and sensitivity offers potential targets for future therapeutic strategies. The findings highlight the importance of personalized treatment approaches and the need for further research to improve radiotherapy outcomes in cancer patients.

## Authors contribution

AV conducted and coordinated in silico studies, and wrote the manuscript; GVM, IVO contributed to the writing of the manuscript; ANO and AA provided helpful suggestions to the experimental design and improvements to the project; AZ and MSK conceived the project; MSK supervised the project, contributed to the writing of the manuscript and is the corresponding author.

## Data availability

All data supporting the conclusions of the paper are available in the article and corresponding figures. TCGA datasets used in the paper are described in the materials and methods section.

